# Genomic and transcriptomic insights into antipsychotic-induced changes in total cholesterol and body mass index in a multi-ancestry cohort of the US veterans

**DOI:** 10.64898/2026.07.20.26358275

**Authors:** Hamed Kazemi, John E. Drake, Silviu-Alin Bacanu, Benjamin H. Mcmahon, Khushbu Agarwal, Alejandro M. Zuniga, Sutanay Choudhury, Hamed Abbaszadegan, Sayera Dhaubhadel, Kory A. Johnson, Julie A. Kreyenbuhl, Stephen R. Marder, Philip D. Harvey, Vladimir I. Vladimirov, Million Veteran Program, MVP Program Office, MVP Steering Committee, MVP Co-Principal Investigators, MVP Core Operations, Ayman H. Fanous

## Abstract

**Objective:** Antipsychotic medications are a cornerstone in the treatment of many psychiatric disorders, but they are associated with adverse metabolic effects including weight gain and hypercholesterolemia. To investigate the underlying genetic architecture of these effects, we conducted the largest-by-far and most ethnically diverse genome-wide association study (GWAS) of longitudinal changes in total cholesterol (ΔTC) and body mass index (ΔBMI) using an antipsychotic-treated cohort from the Million Veteran Program (MVP).

**Methods:** The study included 59,372 participants for ΔTC and 39,112 for ΔBMI across European (EUR), African American (AFR), and Hispanic (HIS) ancestries. GWAS, trans-ancestry meta-analysis, functional annotation, and summary-data-based Mendelian randomization (SMR) analyses were performed to identify associated loci, enriched biological pathways, and gene expression signals.

**Results:** We identified genome-wide significant and suggestive loci for both traits, with stronger associations for ΔTC and clinically significant weight gain (ΔBMI > 1.5 kg/m²) than for overall BMI change. Significant loci included genes involved in cholesterol metabolism and lipid homeostasis for ΔTC and genes previously implicated in BMI-related traits for ΔBMI > 1.5 kg/m². Trans-ancestry meta-analysis highlighted suggestive loci shared across ancestries, and functional annotation demonstrated significant enrichment of protein homeostasis and synaptic function gene sets for ΔBMI > 1.5 kg/m². SMR analyses further identified suggestive gene expression associations in whole blood and liver.

**Conclusions:** These findings implicate lipid metabolism, body weight regulation, and central nervous system mechanisms in antipsychotic-associated metabolic changes. This work advances understanding of genetic susceptibility to metabolic adverse effects of antipsychotic treatment and may inform future precision medicine approaches to risk prediction and treatment selection.

## Introduction

Antipsychotic drugs are associated with metabolic dysfunction, and cause adverse metabolic effects that commonly include weight gain, hypercholesterolemia, and elevated levels of insulin, glucose, total cholesterol (TC), low-density lipoprotein (LDL) cholesterol, and triglycerides (TGs) (1). Antipsychotics can affect cholesterol biosynthesis and lipid metabolism, contributing to obesity and related metabolic disorders (2, 3). Second-generation antipsychotics (SGAs) are more strongly linked to metabolic dysregulation than first-generation antipsychotics (FGAs), and patients with psychiatric disorders and treated with SGAs often exhibit notable metabolic alterations (3, 4). SGAs, particularly clozapine and olanzapine, are associated with greater weight gain, elevated TC and TGs, and hyperlipidemia than other types (5–7).

Plasma lipid levels and body weight are strongly influenced by genetic factors. TC heritability is estimated at 40-60%, while genetics accounts for approximately 40-70% of variations in weight gain and obesity (8, 9). Hypercholesterolemia, characterized by elevated cholesterol levels, is among the most prevalent risk factors for coronary artery disease (CAD), which remains the leading cause of death worldwide (10). Patients with psychiatric disorders are at increased risk of dyslipidemia and obesity, which are associated with greater symptom severity and worse clinical outcomes (6, 11, 12). Genetic studies, including multi-ancestry genome-wide association studies (GWASs), have uncovered numerous loci associated with lipid and body weight-related traits in antipsychotic-treated patients (13–15). Large-scale efforts such as the Global Lipids Genetics Consortium (GLGC) and the Genetic Investigation of ANthropometric Traits Consortium (GIANT) have greatly advanced our understanding of the genetic basis of blood lipid traits and human body size and shape, including height and measures of obesity such as body mass index (BMI) (16, 17). Many lipid-associated loci are also linked to cardiovascular and metabolic conditions including BMI (13). Evidence suggests that loci associated with antipsychotic-induced weight gain may act through lipid pathway dysfunction (18). Furthermore, pharmacogenetic studies have identified loci associated with antipsychotic-induced changes in lipid levels and BMI (19, 20). Collectively, these findings suggest that genetic factors contribute to susceptibility to antipsychotic-induced hyperlipidemia and weight gain, potentially through biological pathways that overlap with those underlying primary obesity and lipid regulation.

The Million Veteran Program (MVP) of the U.S. Department of Veterans Affairs is one of the world’s largest and most diverse biobanks, linking genetic data with extensive electronic health records (EHRs) (21). In this study, we conducted the largest and most ethnically diverse GWAS of longitudinal changes in total cholesterol (ΔTC) and body mass index (ΔBMI) among antipsychotic-treated participants from the MVP, including European (EUR), African American (AFR), and Hispanic (HIS) ancestries, to identify genetic loci associated with antipsychotic-induced metabolic changes and explore their biological relevance.

## Methods

### Study population and phenotypes

We utilized data from the MVP, comprising over 658,000 U.S. veterans. Our analytic cohort included 59,372 patients for TC (EUR = 39,117; AFR = 14,621; HIS = 5,634) and 39,112 for BMI (EUR = 24,751; AFR = 10,661; HIS = 3,700), all prescribed antipsychotic medications. The primary phenotypes of interest were ΔTC and ΔBMI. In addition, a secondary BMI phenotype was defined as ΔBMI > 1.5 kg/m², representing a clinically meaningful increase in BMI from baseline during follow-up. For each patient, we calculated the mean of all measurements obtained within one year before and withing one year after antipsychotic initiation. ΔTC and ΔBMI were then derived as the log_10_-transformed value (22) of the post-treatment mean divided by the pre-treatment mean (ΔTC/ΔBMI = log_10_(post-treatment mean) – log_10_(pre-treatment mean)).

#### Genotyping

Participants in the MVP provided peripheral blood samples for DNA extraction and biobanking at the VA Central Biorepository (Boston, MA). Genotyping was performed using the MVP 1.0 custom Affymetrix Axiom Biobank Array (∼723,000 markers) (21). This array includes exonic variants, disease-associated SNPs, and ancestry-informative markers optimized for diverse populations.

#### Stepwise regression to select covariates

To identify appropriate covariates for inclusion in association analyses, we applied stepwise regression (23) using ΔTC and ΔBMI phenotypes as dependent variables. Covariates included age, age², sex, and ancestry-specific principal components (PCs). PCs were retained to adjust for population stratification, with the number of PCs selected for each ancestry group determined by stepwise regression.

#### GWAS analysis

GWAS analyses were performed using PLINK 2.0. Prior to GWAS, variants with a minor allele frequency below 1% were excluded (*--maf 0.01*). Association testing was carried out with the *--glm firth hide-covar* option. Continuous covariates were standardized with *--covar-variance-standardize* to mean zero and unit variance. Variants with Minimac3 imputation R² > 0.3 in at least two ancestry groups were retained. Finally, collinearity among covariates was minimized by removing pairs with correlations above 0.95 using the *--max-corr 0.95* flag. LocusZoom (24) (http://locuszoom.org/) was used to generate regional association plots for GWAS results.

#### Trans-ancestry Meta analysis

We performed a trans-ancestry meta-analysis of GWAS summary statistics for ΔTC and ΔBMI using METASOFT (25) (http://genetics.cs.ucla.edu/meta/index.html). GWAS summary statistics from EUR, AFR, and HIS ancestry groups were included. Trans-ancestry meta-analysis was conducted using the Han and Eskin random-effects model (RE2) implemented in METASOFT.

#### Functional Mapping and Annotation (FUMA) analysis

FUMA analysis of GWAS meta-analysis results was performed using the FUMA platform v1.5 (https://fuma.ctglab.nl/) (26). Summary statistics from the meta-analyses of ΔTC and ΔBMI were uploaded to FUMA and processed using default parameters. SNPs were mapped to genes using positional mapping (±10 kb). Gene-based association and gene-set enrichment analyses were then performed using MAGMA within FUMA and MSigDB gene sets. Gene-set significance was assessed using MAGMA and Bonferroni correction. Gene sets with Bonferroni-corrected p-values < 0.05 were considered statistically significant, while gene sets with nominal p-values < 0.05 were considered suggestive.

#### Summary-data-based Mendelian Randomization (SMR) analysis

We conducted SMR analysis to investigate potential pleiotropic associations between gene expression and phenotypes using SMR software v1.4.0 (27) (https://yanglab.westlake.edu.cn/software/smr/). GWAS summary statistics for the phenotypes of interest were used as input together with cis-eQTL summary statistics from GTEx v8. SMR analyses were performed using eQTL data derived from whole-blood and liver tissues. To distinguish true pleiotropic or potentially causal associations from those driven by linkage, the HEIDI (Heterogeneity in Dependent Instruments) test was performed for each SMR association.

Associations with SMR P-value below the predefined significance threshold (P_SMR_ ≤ 5 × 10^−8^) and a non-significant HEIDI test (P_HEIDI_ > 0.01) were considered consistent with a pleiotropic or potentially causal relationship rather than linkage (27, 28). Associations meeting the suggestive threshold were defined as P_SMR_ < 0.005 and P_HEIDI_ > 0.01. The HEIDI test was performed only when at least three independent SNPs were available; results were reported as not available (NA) when fewer than three SNPs passed quality-control and filtering steps.

## Results

### GWAS results

Among the genome-wide significant loci identified across the EUR, AFR, and HIS ancestry groups, several genes have previously been implicated in cholesterol and lipid metabolism, as well as in BMI, weight gain, and obesity-related traits. These findings are discussed in greater detail in the Discussion section.

For ΔTC, we observed genome-wide significant associations on chromosome 7 including *ORC5* (29) and *RELN* (30–32), and chromosome 11 containing *EHD1* (33), *ATG2A* (34, 35), and *TM7SF2* (36–39) in the EUR cohort, genes with prior evidence linking them to cholesterol or lipid metabolism. In the AFR cohort, a significant association was detected on chromosome 7 and the *DPP6* gene; however current literature does not support a direct or indirect role for *DPP6* in cholesterol or lipid-related pathways. Suggestive associations were detected on chromosomes 7, 14, and 22 in EUR; chromosomes 2, 12, 13, 16, and 19 in AFR; and chromosome 5 in the HIS cohort. For ΔBMI, no associations reached genome-wide significance. However, suggestive peaks were observed on chromosomes 2, 3, 4, 8, and 12 in the EUR cohort; chromosomes 3, 6, 17, and 22 in the AFR cohort; and chromosomes 7, 8, 12, and 16 in the HIS cohort. For ΔBMI > 1.5 kg/m², genome-wide significant associations were observed on chromosome 8 including *ADAM28* (40, 41), and *ADAMDEC1* (42, 43) in the EUR cohort and on chromosomes 4 and 5 including *NDST3*, *PROP1*, and *COL23A1* (44) in the AFR cohort. Notably, several of these genes have demonstrated direct links to BMI or obesity-related pathways, while *NDST3* and *PROP1* may influence these traits more indirectly. Suggestive associations were found on chromosomes 2, 3, 4, 9, and 11 in EUR; chromosomes 1, 3, 4, 5, 6, 7, 9, and 14 in AFR; and chromosomes 2, 3, 4, 9, 12, and 15 in HIS. Table 1 summarizes the genome-wide significant SNPs identified in our analyses, together with genes located within ±500 kb of each lead SNP for ΔTC and ΔBMI > 1.5 kg/m². Figures 1– 4 present the genome-wide Manhattan plot peaks from GWAS conducted across different ancestries for the studied phenotypes. Complete tables and figures of all suggestive results are provided in Supplemental File 1 (Tables S2, S3, S4) and Supplemental File 2 (Figures S5, S6, S7, S8, S9), respectively.

**Table 1.** Genome-wide significant results of GWAS for ΔTC and ΔBMI > 1.5 kg/m².

| Phenotype | Ethnicity | CHR | SNP | Nearest gene | $-\log_{10}(P)$ | $\pm 500$ kb genes |
| --- | --- | --- | --- | --- | --- | --- |
| ΔTC | EUR | 7 | 7: 103,747,071 A/G | *ORC5* | 7.796 | *RELN*, *ORC5*, *LHFPL3* |
|  |  | 11 | 11: 64,743,730 G/A | *MAJIN* | 7.416 | *SLC22A11*, *SLC22A12*, *NRXN2*, *RASGRP2*, *PYGM*, *SF1*, *MAP4K2*, *MEN1*, *CDC42BPG*, *EHD1*, *ATG2A*, *PPP2R5B*, *GPHA2*, *MAJIN*, *BATF2*, *ARL2*, *SNX15*, *SAC3D1*, *CDCA5*, *ZFPL1*, *VPS51*, *TM7SF2*, *ZNHIT2*, *SPDYC*, *CAPN1*, *POLA2*, *CDC42EP2*, *DPF2*, *TIGD3*, *SLC25A45*, *FRMD8* |
|  | AFR | 7 | 7: 154,056,996 C/T | *DPP6* | 8.510 |  |
| ΔBMI $> 1.5$ kg/m<sup>2</sup> | EUR | 8 | 8: 24,283,237 A/C | *AC120193.1* | 8.063 | *ADAM28*, *ADAMDEC1*, *ADAM7*, *NEFM* |
|  | AFR | 4 | 4: 118,572,022 G/C | *LINC01378* | 7.442 | *NDST3* |
|  |  | 5 | 5: 177,747,691 T/C | *COL23A1* | 7.320 | *PROP1*, *N4BP3*, *RMND5B*, *NHP2*, *GMCL2*, *HNRNPAB*, *PHYKPL*, *COL23A1*, *CLK4*, *AC113348.1*, *AC113348.2*, *ZNF354A* |

**Figure 1.**
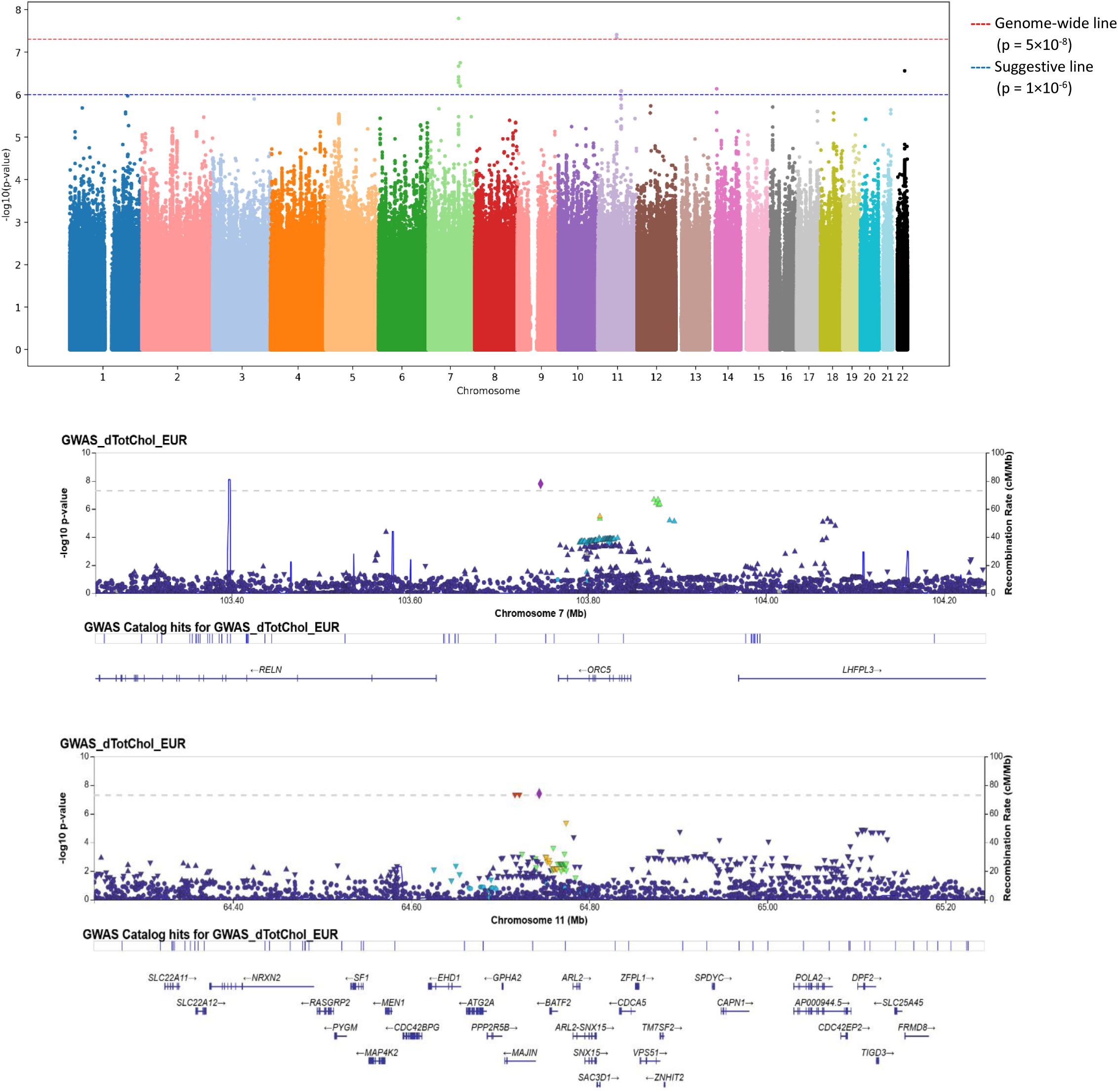
Manhattan plot of GWAS results for ΔTC in EUR cohort

**Figure 2.**
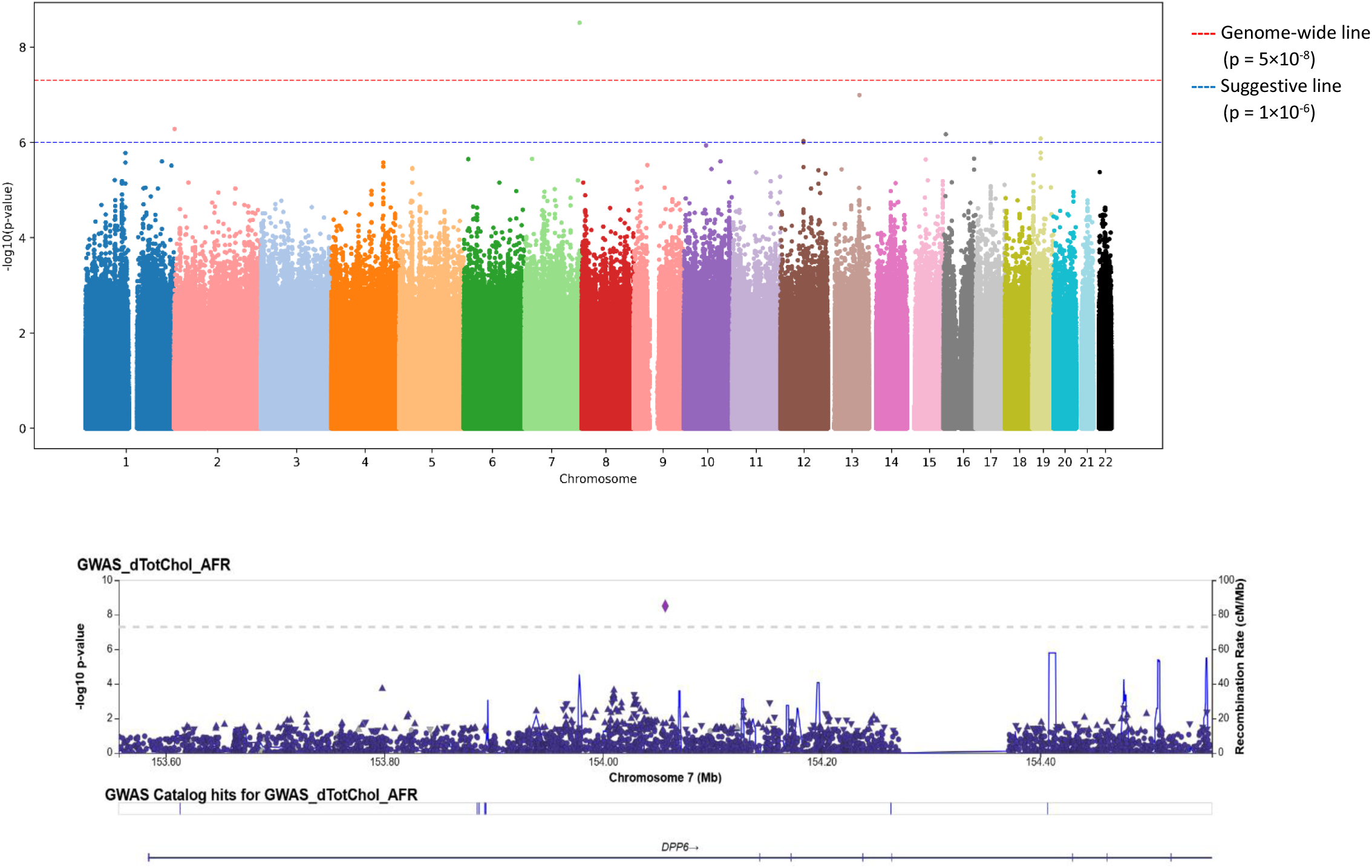
Manhattan plot of GWAS results for ΔTC in AFR cohort

**Figure 3.**
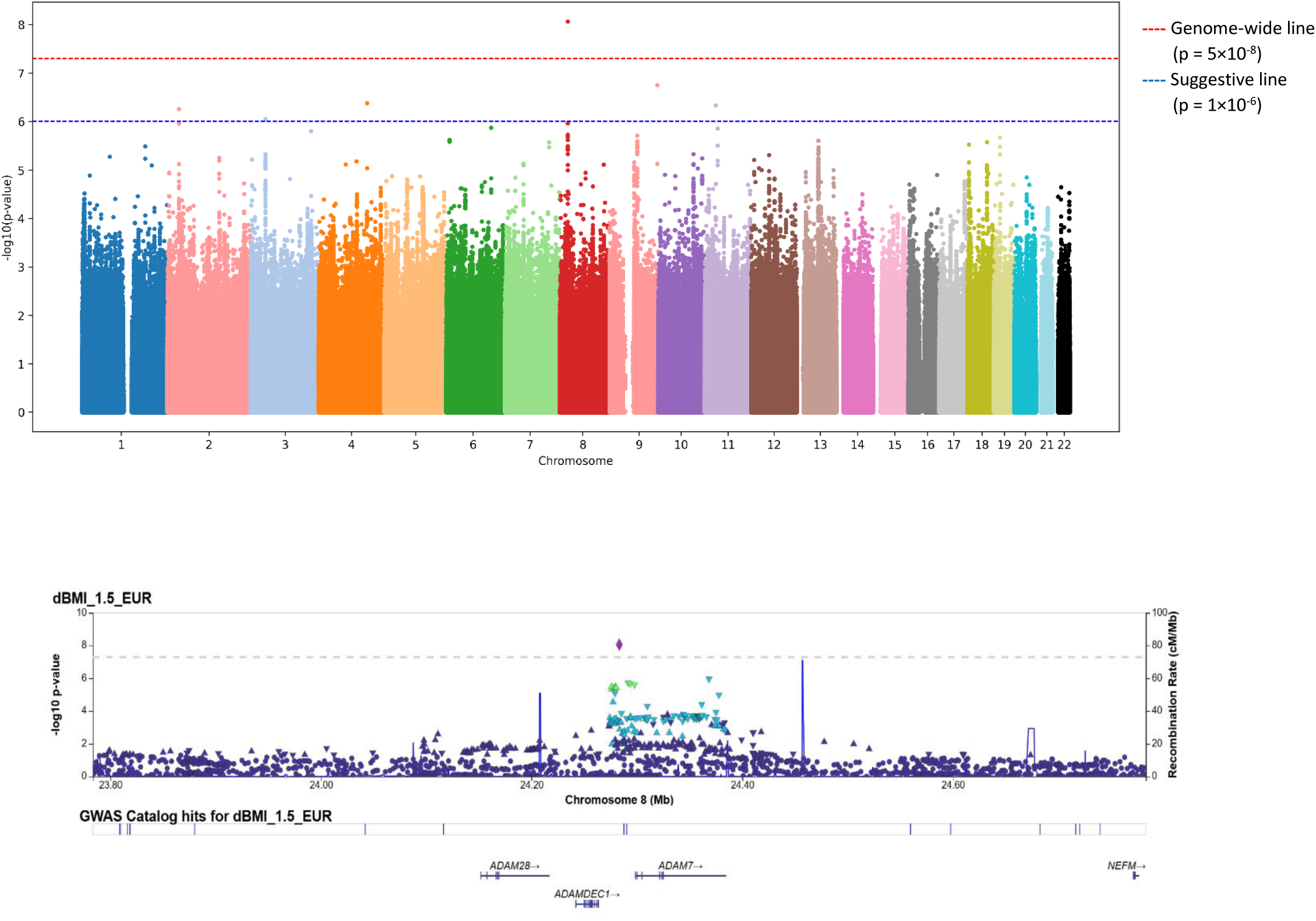
Manhattan plot of GWAS results for ΔBMI > 1.5 kg/m² in EUR cohort

**Figure 4.**
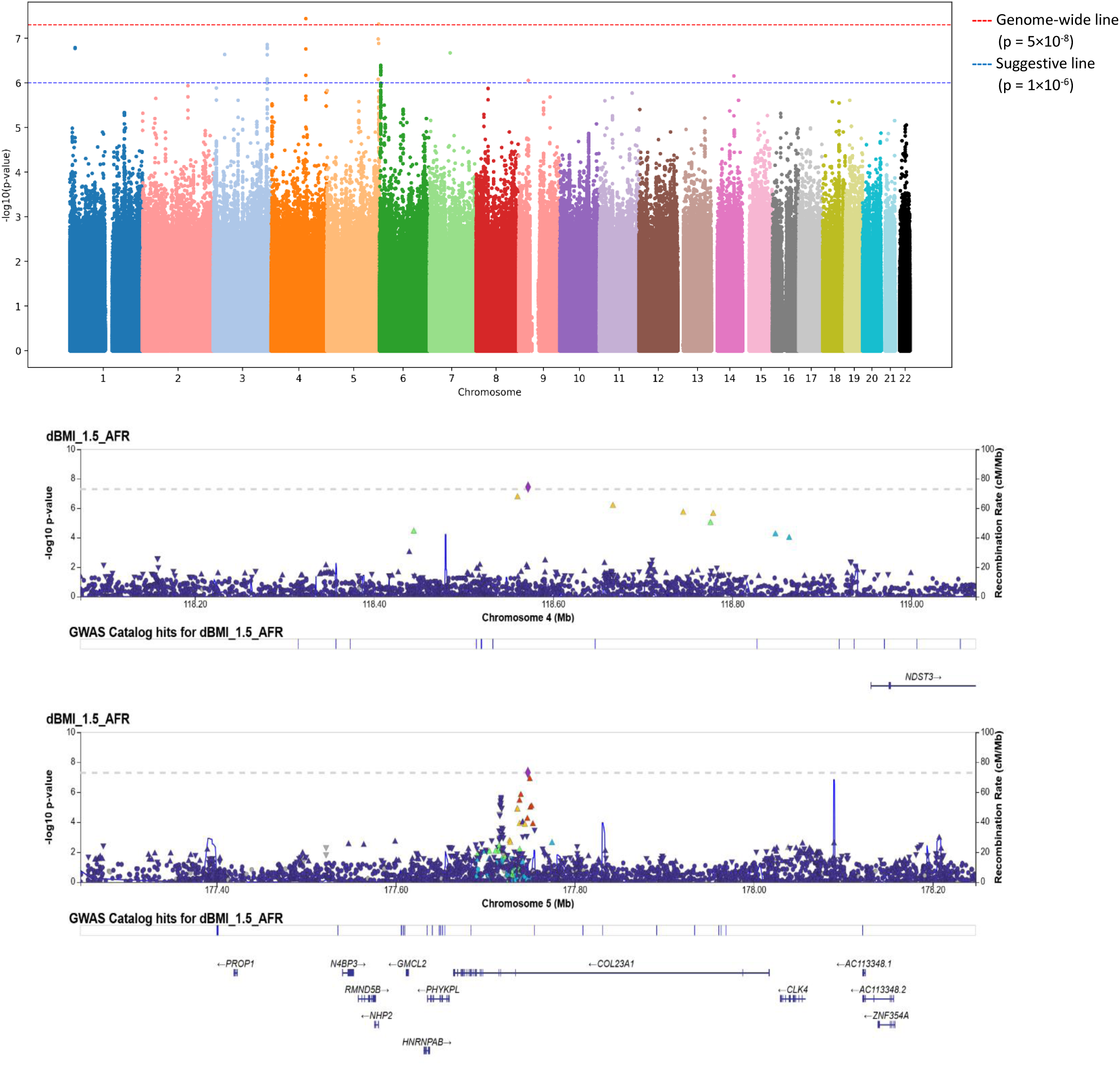
Manhattan plot of GWAS results for ΔBMI > 1.5 kg/m² in AFR cohort

### Meta analysis results

For ΔTC, ΔBMI, and ΔBMI > 1.5 kg/m² no loci reached genome-wide significance in the meta-analysis; however, several suggestive associations were observed across multiple chromosomes, mapping to genes located within ±500 kb of the lead SNP. For ΔTC and ΔBMI > 1.5 kg/m², some genes that achieved genome-wide significance in the ancestry-specific GWAS were also identified in the meta-analysis but did not retain genome-wide significance. For ΔTC, this included *DPP6* on chromosome 7, the top genome-wide locus in the GWAS for AFR, which has not been linked to any cholesterol or lipid metabolism phenotypes to date. For ΔBMI > 1.5 kg/m², implicated genes included *ADAM28* (40, 41) and *ADAMDEC1* (42, 43) on chromosome 8, which represented the top genome-wide loci in the GWAS for EUR. These genes have previously been associated with body weight–related traits. The lead SNPs identified in the trans-ancestry meta-analysis of GWASs across the EUR, AFR, and HIS cohorts, together with all genes located within ±500 kb of each SNP, are provided in Supplemental File 1 (Table S5). The Manhattan plots for the meta-analyses for ΔTC, ΔBMI, and ΔBMI > 1.5 kg/m² are provided in Supplemental File 2 (Figures S10, S11, S12).

### FUMA analysis results

FUMA pathway analysis identified several pathways showing nominal associations with ΔTC and ΔBMI in the meta-analysis (Supplemental File 1; Tables S6, S7). In contrast, for ΔBMI > 1.5 kg/m², a subset of pathways remained significant after Bonferroni correction (Supplemental File 1; Table S8). For ΔTC, FUMA identified nominal enrichment of lipid metabolism-related pathways including steroid biosynthesis and acylglycerol homeostasis, providing biological context for the GWAS findings. For ΔBMI, some nominally enriched pathways were related to neuronal signaling and development, including positive regulation of receptor clustering and negative regulation of axonogenesis. For ΔBMI > 1.5 kg/m², pathways including regulation of protein autoubiquitination and synaptic vesicle localization remained significant after Bonferroni correction, highlighting mechanisms involved in protein homeostasis and synaptic transmission (45). Additional nominally enriched pathways included synaptic vesicle transport, vesicle localization, and regulation of intracellular calcium release, further supporting a role for neuronal signaling in weight gain (46, 47). Collectively, these results highlight neuro-metabolic mechanisms as potential contributors to BMI regulation, consistent with prior GWAS findings for obesity-related traits.

### SMR analysis results

SMR analyses identified only suggestive associations across all three phenotypes. Analyses were restricted to whole-blood and liver eQTL datasets. Associations with a P_SMR_ < 0.005 and P_HEIDI_ > 0.01 were considered suggestive and consistent with a pleiotropic or potentially causal relationship, whereas associations failing the HEIDI test were interpreted as likely reflecting heterogeneity and linkage. The suggestive associations were found between genetically predicted gene expression in whole-blood and ΔTC for genes implicated in cholesterol and lipid metabolism including *SERPINE2* (chr2) and *PRDX6* (chr1) in the EUR cohort, although other signals did not pass the HEIDI test, indicating that their signals may reflect linkage rather than pleiotropy. For ΔBMI, suggestive associations were detected between genetically predicted whole-blood expression of genes previously linked to weight gain and metabolic regulation, obesity, and BMI encompassing *KHK* (chr2) and *NFIL3* (chr9) in the AFR and *CCDC125* (chr5) in the HIS cohorts, as well as liver expression of *SEPT12* (chr16) in the EUR. For ΔBMI > 1.5 kg/m², suggestive associations included *CISD1* (chr10) in the EUR cohort and *CRAT* (chr9) in the AFR cohort. Complete SMR results are provided in Supplemental File 1 (Tables S9, S10, S11, S12, S13, S14), with genes highlighted in red indicating those previously reported to be directly or indirectly associated with TC and/or BMI. Associations that did not pass the HEIDI test (P_HEIDI < 0.01 or NA) are also included.

## Discussion

This study identified several ancestry-specific and shared genetic signals associated with changes in ΔTC and ΔBMI, in the context of antipsychotic use, with stronger genome-wide evidence observed for ΔTC and ΔBMI > 1.5 kg/m², compared to ΔBMI overall.

For ΔTC in the EUR cohort, genome-wide significant signals were identified in genes with emerging links to cholesterol metabolism and lipid homeostasis, including *ORC5*, *RELN*, *TM7SF2*, *ATG2A*, and *EHD1*. These genes converge on cholesterol biosynthesis and regulation of lipid metabolism, supporting the biological relevance of our findings. *ORC5* has been associated with higher LDL-C levels and also indirectly affects lipid metabolism through altering regulatory motifs for *FOXA* (29). *RELN*, which encodes protein Reelin, has been linked to cholesterol metabolism, particularly in the context of atherosclerosis and schizophrenia. Evidence suggests that Reelin contributes to cholesterol efflux (30), while studies in schizophrenia showed decreased *RELN* expression and changes in lipid profiles, including elevated TC (31). In addition, *RELN* may also contribute to lipid metabolic processes in the adult and aging brain (32). *TM7SF2* plays an important role in cholesterol biosynthesis under stress conditions and has been associated with inflammatory responses (36). Prior studies implicate *TM7SF2* in regulating lipid metabolism and reprogramming (37, 38), and dysregulation of *TM7SF2* as one of the cholesterol metabolism genes, may contribute to metabolic abnormalities such as dyslipidemia and obesity (39). *ATG2A* has also been associated with the regulation of lipid droplet morphology and dispersion (34), and promotion of lipid droplet growth through transferring triacylglycerol (35). In parallel, *EHD1* contributes to intracellular cholesterol homeostasis, regulation of lipid droplet formation, and internalization and recycling of LDL receptors, thereby influencing cellular cholesterol uptake and storage (33). The AFR analysis identified a genome-wide significant signal on chromosome 7 overlapping the EUR chromosomal region but mapped to a different gene as *DPP6.* Although *DPP6* has not previously been implicated in lipid or cholesterol-related traits, this might reflect potential ancestry-specific effects or novel biological mechanisms that warrant further investigation and replication. Furthermore, suggestive loci identified in the HIS cohort indicate heterogeneity in genetic architecture of ΔTC across ancestries. Unlike ΔTC, ΔBMI yielded only suggestive associations across ancestries. This pattern may reflect smaller effect sizes, greater polygenicity, and stronger environmental influences on modest changes in body weight. The absence of genome-wide significance for ΔBMI may reflect the heterogeneous nature of moderate weight change compared to extreme phenotypes. In contrast, ΔBMI > 1.5 kg/m² demonstrated stronger and biologically coherent associations in our study, suggesting that substantial weight gain may better capture individuals with increased genetic susceptibility. In the EUR cohort, a genome-wide significant association was identified on chromosome 8 near *ADAM28* and *ADAMDEC1*. In addition, in the AFR cohort, genome-wide significant signals were identified on chromosome 4 near *NDST3* and on chromosome 5 near *PROP1* and *COL23A1*. In human studies, elevated *ADAM28* expression in blood cells has been associated with features of metabolic syndrome, including higher BMI and altered HDL cholesterol levels (40). Furthermore, *ADAM28* knockout mice exhibited reduced body weight, increased HDL cholesterol, and improved insulin sensitivity and glucose tolerance, supporting a functional role for *ADAM28* in metabolic regulation (41). *ADAMDEC1* has also been identified as an inflammation-related gene that is overexpressed in the intestinal mucosa of obese rat models (42). Conversely, loss of *ADAMDEC1* in mice resulted in greater weight loss, increased inflammation, and impaired intestinal immune function, suggesting a protective role for *ADAMDEC1* in metabolic and intestinal homeostasis (43). Among the AFR-specific findings, *NDST3* and *PROP1* have not been directly associated with BMI or body weight-related traits in previous studies. However, both genes have biological functions that could indirectly influence body weight and BMI through central nervous system (controlling appetite and energy balance) and endocrine (metabolism and growth regulation hormones) mechanisms, requiring investigation. Variants in *COL23A1* have been associated with the risk of weight regain after weight loss in a sex-specific manner, potentially demonstrating additive effects on body weight regulation. It has been proposed that *COL23A1* may affect weight regulation by modulating adipocyte–extracellular matrix interactions during weight loss and influencing preadipocyte differentiation (44). Overall, the stronger signals observed for ΔBMI > 1.5 kg/m² support the hypothesis that extreme weight gain phenotypes may enrich for greater genetic susceptibility, consistent with threshold or extreme-phenotype models in complex trait genetics.

The meta-analyses identified suggestive associations near several genes that were also detected in the ancestry-specific GWAS analyses, including *DPP6* for ΔTC, and *ADAM28* and *ADAMDEC1* for ΔBMI > 1.5 kg/m². Notably, all these loci reached genome-wide significance in individual ancestry-specific analyses, further supporting the consistency of these signals across populations. Although *DPP6* has not been implicated in cholesterol or lipid metabolism, its recurrence in ancestry-specific GWAS and meta-analysis raises the possibility of a novel or ancestry-specific contribution to ΔTC variation, requiring further validation. In addition, the repeated identification of *ADAM28* and *ADAMDEC1* across analyses provides additional support for their potential involvement in weight gain regulation and BMI phenotype.

FUMA pathway analysis identified significant enrichment of regulation of protein autoubiquitination and synaptic vesicle localization, suggesting roles for the ubiquitin–proteasome system and synaptic function in metabolic regulation and obesity susceptibility. Evidence supports the involvement of ubiquitin-related pathways in energy balance and metabolic homeostasis.

Overexpression of *Ubqln1* in transgenic mice reduced body weight gain, visceral fat, leptin and insulin levels while upregulating energy-sensing proteins across multiple tissues and elevating metabolic rate (48). In humans, plasma ubiquitin levels are decreased in obesity and inversely correlated with BMI (49). Furthermore, impaired hypothalamic ubiquitin–proteasome function has been linked to obesity through disruption of protein turnover and energy homeostasis, whereas preservation of this pathway appears protective against weight gain (50). The enrichment of synaptic vesicle localization pathway supports a role for central nervous system mechanisms in obesity-related phenotypes, consistent with large-scale BMI GWAS meta-analyses from the GIANT Consortium that highlighted pathways involving synaptic function, glutamatergic signaling, insulin action, energy metabolism, and adipogenesis (46). The synaptic vesicle localization pathway is enriched in brain-related functions and plays central role in neuronal signaling and synaptic function (51). In addition, synaptic processes, especially glutamatergic and GABAergic neurotransmission, are critical for the regulation of energy balance, and synaptic dysfunction has been proposed to contribute to metabolic dysregulation and obesity (52).

Altogether, genes within the regulation of protein autoubiquitination and synaptic vesicle localization pathways are not individually identified as genome-wide significant BMI loci, their enrichment for neuronal and synaptic processes that have been consistently implicated in large-scale studies (GIANT Consortium) (46), supports a pathway-level contribution of central nervous system mechanisms (neuronal and synaptic) to obesity susceptibility.

Further SMR analyses provided functional context by identifying suggestive associations between genetically predicted gene expression in whole blood and liver and both ΔTC and ΔBMI phenotypes. Although several genes with reported links to lipid metabolism, adiposity, energy homeostasis, body weight regulation, and metabolic disorders were identified (*SERPINE2* and *PRDX6* for ΔTC, and *CISD1*, *KHK*, *NFIL3*, *CRAT*, *CCDC125*, and *SEPT12* for ΔBMI-related phenotypes), all associations remained suggestive and some associations did not pass HEIDI filtering, indicating that observed signals may reflect linkage rather than shared causal variants and should be interpreted cautiously pending independent replication.

Across phenotypes and ancestries, our findings revealed convergent biological themes involving cholesterol biosynthesis and lipid metabolism, adiposity and obesity regulation, endocrine and metabolic signaling, and central nervous system mechanisms related to appetite and energy balance. While ΔTC loci were predominantly enriched for genes directly involved in cholesterol homeostasis and lipid handling, ΔBMI and particularly ΔBMI > 1.5 kg/m² implicated broader networks related to adiposity, neuronal regulation of food intake, endocrine control, metabolic regulation, and inflammation. SMR analyses further supported these findings by highlighting genes nominally associated with lipid levels and metabolism, dyslipidemia, adipose tissue regulation, BMI and body weight regulation, obesity, and metabolic dysfunction across ancestries in whole blood and liver tissues. Notably, stronger and biologically coherent signals were observed for ΔBMI > 1.5 kg/m² compared to modest BMI change, supporting that substantial weight gain may better capture genetically driven susceptibility. In addition, FUMA identified enrichment of gene sets involving protein homeostasis and synaptic function, reinforcing evidence that protein turnover and brain-enriched synaptic mechanisms contribute to obesity susceptibility and energy homeostasis. Together, this multi-ancestry GWAS combined with FUMA and integrative SMR analyses provides new insights into the genetic architecture of longitudinal changes in TC and BMI in the context of antipsychotic use. However, smaller non-European ancestry samples, heterogeneity in antipsychotic exposure and clinical characteristics, and the absence of independent replication are important limitations of this study. These findings advance our understanding of the biological mechanisms influencing lipid regulation and body weight dynamics and underscore the importance of integrative multi-ancestry genomic approaches in uncovering the genetic basis of metabolic changes.

## Supporting information

Supplemental Appendix for VA Million Veteran Program Core Acknowledgements for Publications

## Data Availability

Million Veteran Program data are not publicly available because of privacy and data-security requirements. Eligible investigators may apply for access through the established MVP data-access process, subject to scientific and regulatory approval.

## Acknowledgments

This research is based on data from the Million Veteran Program, Office of Research and Development, Veterans Health Administration, and was supported by MVP068. This publication does not represent the views of the Department of Veteran Affairs or the United States Government. The authors would like to acknowledge the research personnel involved in the Million Veteran Program (see Supplemental Appendix for VA Million Veteran Program Core Acknowledgements for Publications), whose participation and dedication made this study possible. The authors also thank the VINCI and GENISIS support teams and the Million Veteran Program Core Statistical Analysis team for their contributions.

