## Supplemental File 1 for "Genomic and transcriptomic insights into antipsychotic-induced changes in total cholesterol and body mass index in a multi-ancestry cohort of the US veterans"

Table S2. GWAS results for ΔTC (genome-wide significant signals are highlighted in red)

| Phenotype | Ethnicity | CHR | SNP | Nearest gene | -log10(P) | ±500 kb genes |
| --- | --- | --- | --- | --- | --- | --- |
| ΔTC | EUR | 7 | 7: 103,747,071 A/G | ***ORC5*** | 7.796 | ***RELN****, ORC5, LHFPL3* |
|  |  | 11 | 11: 64,743,730 G/A | *MAJIN* | 7.416 | *SLC22A11, SLC22A12, NRXN2, RASGRP2, PYGM, SF1, MAP4K2, MEN1, CDC42BPG,* ***EHD1****,* ***ATG2A****, PPP2R5B, GPHA2, MAJIN, BATF2, ARL2, SNX15, SAC3D1, CDCA5, ZFPL1, VPS51,* ***TM7SF2****, ZNHIT2, SPDYC, CAPN1, POLA2, CDC42EP2, DPF2, TIGD3, SLC25A45, FRMD8* |
|  |  | 7 | 7: 109,674,134 C/G | *RPL3P8* | 6.752 |  |
|  |  | 22 | 22: 41,563,819 T/G | *EP300* | 6.563 | *MCHR1, SLC25A17, ST13, DNAJB7, XPNPEP3, RBX1, EP300, L3MBTL2, CHADL, RANGAP1, ZC3H7B, TEF, TOB3, PHF5A, ACO2, POLR3H, CSDC2, PMM1, DESI1, XRCC6* |
|  |  | 14 | 14: 24,751,063 C/G | *AL096870*.8 | 6.139 | *DHRS4, DHRS4L2, CARMIL3, CPNE6, NRL, PCK2, DCAF11, FITM1, EMC9, RNF31, IRF9, IPO4, TSSK4, CHMP4A, MDP1, NEDD8, TINF2, DHRS1, NOP9, CIDEB, LTB4R2, LTB4R, NFATC4, NYNRIN, CBLN3, KHNYN, SDR39U1, CMA1, CTSG, GZMH, GZMB* |
|  |  | 11 | 11: 80,884,025 G/C | *AP003464.1* | 6.090 |  |
|  | AFR | 7 | 7: 154,056,996 C/T | *DPP6* | 8.510 |  |
|  |  | 13 | 13: 88,122,974 T/TA | *MIR4500HG* | 6.995 | *SLITRK5* |
|  |  | 2 | 2: 823,988 G/T | *LINC01115* | 6.281 | *TMEM18, SNTG2* |
|  |  | 16 | 16: 6,215,788 AT/A | *RBFOX1* | 6.172 |  |
|  |  | 19 | 19: 22,729,122 A/G | *RNU6-1179P* | 6.079 | *ZNF257, ZNF676, ZNF729, ZNF98, ZNF492, ZNF99, ZNF723, ZNF728* |
|  |  | 12 | 12: 65,225,529 A/G | *TBC1D30* | 6.028 | *C12orf56, XPOT, TBK1, RASSF3, GNS, TBC1D30, AC078815.1, WIF1, LEMD3, MSRB3* |
|  | HIS | 5 | 5: 94,899,743 G/A | *ARSK* | 6.752 | *MCTP1, FAM81B, TTC37, ARSK, GPR150, RFESD, SPATA9, RHOBTB3, GLRX, ELL2* |

Table S3. GWAS results for ΔBMI

| Phenotype | Ethnicity | CHR | SNP | Nearest gene | -log10(P) | ±500 kb genes |
| --- | --- | --- | --- | --- | --- | --- |
| ΔBMI | EUR | 12 | 12: 59,897,323 C/G | *LINC02448* | 6.938 | *SLC16A7* |
|  |  | 8 | 8: 140,870,200 T/G | *TRAPPC9* | 6.312 | *KCNK9, TRAPPC9* |
|  |  | 3 | 3: 11,526,888 T/G | *ATG7* | 6.263 | *SLC6A1, HRH1, ATG7, VGLL4, TAMM41* |
|  |  | 2 | 2: 33,881,983 G/A | *AC017050.1* | 6.226 | *LTBP1, RASGRP3, FAM98A* |
|  |  | 4 | 4: 69,338,138 AAATT/A | *TMPRSS11E* | 6.137 | *TMPRSS11F, TMPRSS11B, YTHDC1, TMPRSS11E, UGT2B17, UGT2B15, UGT2B10, UGT2A3* |
|  | AFR | 22 | 22: 17,529,434 CAT/C | *CECR7* | 6.585 | *CCT8L2, XKR3, GAB4, IL17RA, CECR7, TMEM121B, HDHD5, ADA2, CECR2* |
|  |  | 17 | 17: 36,477,179 C/T | *MRPL45* | 6.576 | *DDX52, HNF1B, TBC1D3K, TBC1D3F, TBC1D3L, TBC1D3, MRPL45, GPR179, SOCS7, ARHGAP23, SRCIN1, EPOP, MLLT6, RNA5SP440, CISD3, PCGF2, PSMB3, CWC25* |
|  |  | 3 | 3: 34,044,916 G/A | *LINC01811* | 6.340 | *CLASP2, PDCD6IP* |
|  |  | 3 | 3: 84,765,690 C/G | *LINC00971* | 6.272 | *CADM2* |
|  |  | 6 | 6: 158,553,908 A/G | *SERAC1* | 6.269 | *ZDHHC14, SNX9, SYNJ2, SERAC1, GTF2H5, TULP4, TMEM181* |
|  | HIS | 7 | 7: 136,859,575 T/C | *AC009264.1* | 6.505 | *CHRM2, PTN, DGKI* |
|  |  | 16 | 16: 15,230,638 G/A | *PDXDC1, PKD1P6* | 6.427 | *BFAR, PLA2G10, NPIPA3, NPIPA2, NOMO1, NPIPA1, PDXDC1, PKD1P6, NTAN1, RRN3, NPIPA5, MPV17L, AC140504.1, BMERB1, MARF1* |
|  |  | 12 | 12: 92,825,506 C/T | *AC063949.2* | 6.158 | *PLEKHG7, EEA1* |
|  |  | 8 | 8: 58,578,300 C/T | *AC104051.1* | 6.147 | *FAM110B* |
|  |  | 16 | 16: 56,894,505 G/T | *MIR138-2* | 6.039 | *AMFR, NUDT21, OGFOD1, BBS2, MT4, MT3, MT2A, MT1E, MT1M, MT1A, MT1B, MT1G, NUP93, SLC12A3, HERPUD1, CETP, NLRC5, CPNE2, PSME3IP1, RSPRY1, ARL2BP, PLLP, CCL22* |
|  |  | 8 | 8: 66,387,409 T/C | *LINC01299* | 6.021 | *ARMC1, MTFR1, PDE7A* |

Table S4. GWAS results for ΔBMI > 1.5 kg/m² (genome-wide significant signals are highlighted in red)

| Phenotype | Ethnicity | CHR | SNP | Nearest gene | -log10(P) | ±500 kb genes |
| --- | --- | --- | --- | --- | --- | --- |
| ΔBMI > 1.5 kg/m² | EUR | 8 | 8: 24,283,237 A/C | *AC120193.1* | 8.063 | ***ADAM28****,* ***ADAMDEC1****, ADAM7, NEFM* |
|  |  | 9 | 9: 138,150,226 C/T | *AL390778.2* | 6.750 | *COL5A1, FCN2, FCN1, OLFM1, PPP1R26, C9orf116, MRPS2, LCN1, OBP2A, PAEP, AL354761.1, GLT6D1, LCN9, SOHLH1, KCNT1* |
|  |  | 4 | 4: 140,587,985 G/A | *MGST2* | 6.377 | *ELF2, MGARP, NDUFC1, NAA15, RAB33B, SETD7, MGST2, MAML3* |
|  |  | 11 | 11: 32,533,478 T/C | ***WT1****-AS* | 6.331 | *AL035078.4, RCN1,* ***WT1****, EIF3M, CCDC73, PRRG4, QSER1* |
|  |  | 2 | 2: 33,789,068 C/G | *RASGRP3* | 6.255 | *LTBP1, RASGRP3, FAM98A* |
|  |  | 3 | 3: 42,592,360 CTT/C | *SEC22C* | 6.053 | *TRAK1, CCK, LYZL4, VIPR1, SEC22C, SS18L2, NKTR, ZBTB47, KLHL40, HHATL, CCDC13, AC006059.2, HIGD1A, ACKR2, KRBOX1, AC092042.3, CYP8B1, ZNF662, GASK1A* |
|  | AFR | 4 | 4: 118,572,022 G/C | *LINC01378* | 7.442 | ***NDST3*** |
|  |  | 5 | 5: 177,747,691 T/C | ***COL23A1*** | 7.320 | ***PROP1****, N4BP3, RMND5B, NHP2, GMCL2, HNRNPAB, PHYKPL, COL23A1, CLK4, AC113348.1, AC113348.2, ZNF354A* |
|  |  | 5 | 5: 176,129,310 G/A | *AC113391.1* | 6.984 | *SIMC1, KIAA1191, ARL10, NOP16, HIGD2A, CLTB, FAF2, RNF44, CDHR2, GPRIN1, SNCB, EIF4E1B, TSPAN17, UNC5A, HK3, UIMC1, ZNF346, FGFR4, NSD1* |
|  |  | 3 | 3: 184,164,264 G/T | *LINC02054* | 6.861 | *ABCC5, HTR3D, HTR3C, HTR3F, EIF2B5, DVL3, AP2M1, ABCF3, VWA5B2, ALG3, EEF1AKMT4, ECE2, EIF4G1, FAM131A, POLR2H, CLCN2, THPO, EPHB3, MAGEF1, VPS8* |
|  |  | 1 | 1: 18,288,001 C/G | *AL357509.1* | 6.792 | *ARHGEF10L, ACTL8, IGSF21* |
|  |  | 7 | 7: 70,285,607 C/CATAT | *AC073873.1* | 6.676 | *AUTS2, GALNT17* |
|  |  | 3 | 3: 38,159,641 C/T | *DLEC1, ACAA1* | 6.640 | *ITGA9, CTDSPL, VILL, PLCD1, DLEC1, ACAA1, MYD88, OXSR1, SLC22A13, SLC22A14, XYLB, ACVR2B, EXOG, SCN5A* |
|  |  | 6 | 6: 3,672,435 A/T | *AL033523.1* | 6.395 | *TUBB2B, PSMG4, SLC22A23, PXDC1, FAM50B, PRPF4B, FAM217A, C6orf201, ECI2* |
|  |  | 14 | 14: 77,056,356 C/T | *AC008050.1* | 6.156 | *GPATCH2L, ESRRB, VASH1, ANGEL1, LRRC74A, IRF2BPL* |
|  |  | 9 | 9: 32,879,680 G/A | *APTX* | 6.058 | *ACO1, DDX58, TOPORS, SMIM27, NDUFB6, TAF1L, TMEM215, APTX, DNAJA1, SMU1, B4GALT1, SPINK4, BAG1, CHMP5, NFX1* |
|  | HIS | 4 | 4: 185,031,000 C/CGT | *ENPP6* | 6.780 | *RWDD4, TRAPPC11, STOX2, ENPP6, IRF2* |
|  |  | 2 | 2: 25,362,584 G/A | *EFR3B* | 6.689 | *NCOA1, PTRHD1, CENPO, ADCY3, DNAJC27, EFR3B, POMC, DNMT3A, DTNB* |
|  |  | 12 | 12: 92,798,633 G/A | *CLLU1OS* | 6.402 | *BTG1, PLEKHG7, CLLU1OS, EEA1* |
|  |  | 9 | 9: 131,992,020 G/A | *AL158151.1* | 6.284 | *ZER1, TBC1D13, ENDOG, SPOUT1, AL441992.2, KYAT1, LRRC8A, PHYHD1, AL672142.1, DOLK, NUP188, SH3GLB2, MIGA2, DOLPP1, CRAT, PTPA, AL158151.3, IER5L, C9orf50, NTMT1, ASB6, PRRX2* |
|  |  | 4 | 4: 4,305,114 G/A | *ZBTB49* | 6.273 | *OTOP1, TMEM128, LYAR, ZBTB49, NSG1, STX18* |
|  |  | 3 | 3: 104,804,317 G/T | *ALCAM* | 6.130 |  |
|  |  | 2 | 2: 242,133,878 G/A | *ANO7* | 6.126 | *AQP12A, KIF1A, AGXT, MAB21L4, CROCC2, SNED1, MTERF4, PASK, PPP1R7, ANO7, HDLBP, SEPTIN2, FARP2, STK25, BOK, THAP4, ATG4B, DTYMK, ING5* |
|  |  | 15 | 15: 82,917,624 C/T | *RP13-996F3.5* | 6.062 | *EFL1, SAXO2, GOLGA6L10, GOLGA6L9, RP13-996F3.5, RP13-996F3.4, RPS17, AC245033.1, CPEB1, AP3B2* |

Table S5. Trans-ancestry meta-analysis of GWAS results

| Phenotype | CHR | SNP | P-value EUR | P-value AFR | P-value HIS | M-value EUR | M-value AFR | M-value HIS | Pvalue_RE2 | -log10(P) | ±500 kb genes |
| --- | --- | --- | --- | --- | --- | --- | --- | --- | --- | --- | --- |
| ΔTC | 7 | 7:154056996:C:T | 0.444406 | 3.02E-09 | 0.046195 | 0 | 1 | 0.136 | 1.28E-07 | 6.89 | ***DPP6*** |
|  | 5 | 5:36071497:G:A | 0.0594996 | 3.46E-06 | 0.125878 | 0.865 | 1 | 0.868 | 6.14E-07 | 6.21 | *SPEF2, IL7R, CAPSL, UGT3A1, UGT3A2, LMBRD2, SKP2, NADK2, RANBP3L* |
|  | 5 | 5:36069761:C:G | 0.0563775 | 3.55E-06 | 0.134839 | 0.865 | 1 | 0.864 | 6.48E-07 | 6.19 |  |
| ΔBMI | 12 | 12:128057889:A:G | 0.0015597 | 0.00011181 | 0.215462 | 0.996 | 1 | 0.859 | 3.82E-07 | 6.42 |  |
| ΔBMI > 1.5 kg/m² | 8 | 8:24283237:A:C | 8.64E-09 | 0.0560073 | 0.531175 | 1 | 0 | 0.295 | 9.68E-08 | 7.01 | ***ADAM28****,* ***ADAMDEC1****, ADAM7, NEFM* |
|  | 4 | 4:140587985:G:A | 4.20E-07 | 0.0113352 | 0.073003 | 1 | 0 | 0.824 | 3.84E-07 | 6.42 | *ELF2, MGARP, NDUFC1, NAA15, RAB33B, SETD7, MGST2, MAML3* |
|  | 11 | 11:32533478:T:C | 4.67E-07 | 0.636646 | 0.184075 | 1 | 0.654 | 0.79 | 4.29E-07 | 6.37 | *RCN1, WT1, EIF3M, CCDC73, PRRG4, QSER1* |

Table S6. FUMA gene-set enrichment results for ΔTC

| Gene Set | N. genes | Beta | Beta STD | SE | P | Bonferroni |
| --- | --- | --- | --- | --- | --- | --- |
| KEGG_STEROID_BIOSYNTHESIS | 15 | 0.90192 | 0.025295 | 0.21307 | 1.16E-05 | 0.197168 |
| GOBP_ACYLGLYCEROL_HOMEOSTASIS | 35 | 0.48347 | 0.020701 | 0.12517 | 5.63E-05 | 0.957482 |
| GOBP_SULFUR_COMPOUND_METABOLIC_PROCESS | 296 | 0.1734 | 0.021443 | 0.04524 | 6.35E-05 | 1 |
| GOBP_REGULATION_OF_TELOMERE_CAPPING | 26 | 0.58427 | 0.021567 | 0.15573 | 8.81E-05 | 1 |
| GOBP_POSITIVE_REGULATION_OF_AMYLOID_BETA_CLEARANCE | 8 | 1.1048 | 0.022632 | 0.30308 | 0.000134 | 1 |
| GOBP_DETECTION_OF_MECHANICAL_STIMULUS | 50 | 0.41097 | 0.021024 | 0.11529 | 0.000183 | 1 |
| GOCC_ATG1_ULK1_KINASE_COMPLEX | 6 | 1.1901 | 0.021113 | 0.33433 | 0.000186 | 1 |
| GOBP_MONONUCLEAR_CELL_MIGRATION | 182 | 0.21446 | 0.020858 | 0.061851 | 0.000263 | 1 |
| GOBP_AMYLOID_PRECURSOR_PROTEIN_BIOSYNTHETIC_PROCESS | 12 | 0.77612 | 0.01947 | 0.22406 | 0.000267 | 1 |
| GOBP_TELOMERE_MAINTENANCE_VIA_TELOMERE_LENGTHENING | 78 | 0.30834 | 0.019687 | 0.09113 | 0.000359 | 1 |

Table S7. FUMA gene-set enrichment results for ΔBMI

| Gene Set | N. genes | Beta | Beta STD | SE | P | Bonferroni |
| --- | --- | --- | --- | --- | --- | --- |
| GOMF_PROTEIN_ARGININE_OMEGA_N_ASYMMETRIC_METHYLTRANSFERASE_ACTIVITY | 7 | 1.1748 | 0.022619 | 0.29575 | 3.58E-05 | 0.60814 |
| GOBP_POSITIVE_REGULATION_OF_MAINTENANCE_OF_SISTER_CHROMATID_COHESION | 5 | 1.4425 | 0.023474 | 0.37507 | 6.02E-05 | 1 |
| GOBP_POSITIVE_REGULATION_OF_RECEPTOR_CLUSTERING | 6 | 1.002 | 0.017862 | 0.26059 | 6.05E-05 | 1 |
| GOCC_RIBONUCLEOPROTEIN_GRANULE | 256 | 0.18433 | 0.02132 | 0.048667 | 7.63E-05 | 1 |
| GOBP_OOCYTE_DIFFERENTIATION | 55 | 0.36874 | 0.019875 | 0.10115 | 0.000134 | 1 |
| GOMF_NUCLEOSIDE_DIPHOSPHATE_KINASE_ACTIVITY | 17 | 0.70856 | 0.021254 | 0.19443 | 0.000135 | 1 |
| GOBP_NEGATIVE_REGULATION_OF_AXONOGENESIS | 59 | 0.37801 | 0.021101 | 0.10387 | 0.000137 | 1 |
| GOMF_HISTONE_ARGININE_N_METHYLTRANSFERASE_ACTIVITY | 8 | 0.95364 | 0.019628 | 0.26385 | 0.000151 | 1 |
| LEE_AGING_MUSCLE_DN | 39 | 0.42851 | 0.019458 | 0.11864 | 0.000153 | 1 |
| GOMF_PROTEIN_ARGININE_N_METHYLTRANSFERASE_ACTIVITY | 12 | 0.80764 | 0.020357 | 0.22411 | 0.000157 | 1 |

Table S8. FUMA gene-set enrichment results for ΔBMI > 1.5 kg/m²

| Gene Set | N. genes | Beta | Beta STD | SE | P | Bonferroni |
| --- | --- | --- | --- | --- | --- | --- |
| **GOBP_REGULATION_OF_PROTEIN_AUTOUBIQUITINATION** | 5 | 1.4757 | 0.02412 | 0.29928 | 4.13E-07 | **0.007028** |
| **GOBP_SYNAPTIC_VESICLE_LOCALIZATION** | 52 | 0.46735 | 0.024603 | 0.10183 | 2.24E-06 | **0.03805** |
| GOBP_SYNAPTIC_VESICLE_TRANSPORT | 40 | 0.497 | 0.022955 | 0.11193 | 4.52E-06 | 0.076845 |
| GOBP_VESICLE_LOCALIZATION | 205 | 0.23312 | 0.024267 | 0.052857 | 5.19E-06 | 0.088287 |
| REACTOME_FBXW7_MUTANTS_AND_NOTCH1_IN_CANCER | 5 | 1.3715 | 0.022416 | 0.33555 | 2.19E-05 | 0.372915 |
| GOBP_REGULATION_OF_INOSITOL_1_4_5_TRISPHOSPHATE_SENSITIVE_ CALCIUM_RELEASE_CHANNEL_ACTIVITY | 4 | 1.5465 | 0.02261 | 0.38398 | 2.83E-05 | 0.481134 |
| MIKKELSEN_IPS_ICP_WITH_H3K4ME3_AND_H327ME3 | 118 | 0.29508 | 0.023359 | 0.074949 | 4.14E-05 | 0.704172 |
| GOBP_HISTONE_H2B_UBIQUITINATION | 10 | 0.89724 | 0.020737 | 0.23042 | 4.95E-05 | 0.841345 |
| GOMF_NEDD8_TRANSFERASE_ACTIVITY | 7 | 1.0041 | 0.019417 | 0.26831 | 9.15E-05 | 1 |
| GOBP_REGULATION_OF_CANONICAL_WNT_SIGNALING_PATHWAY | 232 | 0.19599 | 0.021688 | 0.053601 | 0.000128 | 1 |

Table S9. SMR results for ΔTC and blood tissue

| **Ethnicity** | **Ensembl_Gene_ID** | **Chr** | **Gene** | **Top_SNP** | **p_SMR** | **p_HEIDI** | **nSNP_HEIDI** |
| --- | --- | --- | --- | --- | --- | --- | --- |
| EUR | ENSG00000100395 | 22 | *L3MBTL2* | rs9611497 | 7.28E-04 | *NA* | *NA* |
|  | ENSG00000135919 | 2 | *SERPINE2* | rs282271 | 8.76E-04 | 9.95E-01 | 4 |
|  | ENSG00000205534 | 16 | *RP11-345J4.8* | rs2161671 | 1.77E-03 | 7.91E-01 | 4 |
|  | ENSG00000176473 | 14 | *WDR25* | rs7148488 | 2.81E-03 | 2.83E-01 | 9 |
|  | ENSG00000117592 | 1 | *PRDX6* | rs3934575 | 3.82E-03 | 8.23E-01 | 4 |
|  | ENSG00000173852 | 7 | *DPY19L1* | rs1637668 | 4.47E-03 | 5.14E-01 | 3 |
| AFR | ENSG00000108384 | 17 | *RAD51C* | rs2680688 | 4.61E-03 | 1.41E-01 | 5 |
| HIS | ENSG00000139977 | 14 | *NAA30* | rs8005987 | 1.29E-03 | 7.52E-02 | 4 |
|  | ENSG00000214425 | 17 | *LRRC37A4P* | rs242559 | 2.62E-03 | 2.58E-01 | 13 |
|  | ENSG00000112531 | 6 | *QKI* | rs2759386 | 2.63E-03 | *NA* | *NA* |
|  | ENSG00000263503 | 17 | *RP11-707O23.5* | rs242559 | 2.67E-03 | 2.42E-01 | 13 |
|  | ENSG00000264070 | 17 | *DND1P1* | rs242559 | 2.70E-03 | 2.36E-01 | 13 |
|  | ENSG00000204650 | 17 | *CRHR1-IT1* | rs242559 | 2.75E-03 | 2.63E-01 | 13 |
|  | ENSG00000183665 | 8 | *TRMT12* | rs3812474 | 2.88E-03 | *NA* | *NA* |
|  | ENSG00000060982 | 12 | *BCAT1* | rs2291894 | 4.23E-03 | *NA* | *NA* |
|  | ENSG00000196511 | 7 | *TPK1* | rs10231147 | 4.29E-03 | 4.23E-02 | 4 |
|  | ENSG00000197142 | 10 | *ACSL5* | rs2306159 | 4.84E-03 | 3.55E-03 | 8 |

Table S10. SMR results for ΔTC and liver tissue

| **Ethnicity** | **Ensembl_Gene_ID** | **Chr** | **Gene** | **Top_SNP** | **p_SMR** | **p_HEIDI** | **nSNP_HEIDI** |
| --- | --- | --- | --- | --- | --- | --- | --- |
| EUR | ENSG00000100395 | 22 | *L3MBTL2* | rs4822011 | 6.02E-04 | 3.16E-02 | 5 |
|  | ENSG00000253678 | 8 | *RP11-981G7.3* | rs13279329 | 4.05E-03 | 8.38E-01 | 5 |
| HIS | ENSG00000264070 | 17 | *DND1P1* | rs242559 | 3.26E-03 | 1.38E-01 | 9 |
|  | ENSG00000263503 | 17 | *RP11-707O23.5* | rs242559 | 3.31E-03 | 1.01E-01 | 7 |
|  | ENSG00000262881 | 17 | *RP11-669E14.4* | rs242559 | 3.66E-03 | 3.51E-02 | 8 |
|  | ENSG00000186453 | 2 | *FAM228A* | rs4665253 | 3.89E-03 | 4.16E-01 | 9 |
|  | ENSG00000243384 | 3 | *RP11-475O23.2* | rs6776877 | 4.22E-03 | 9.70E-01 | 3 |

Table S11. SMR results for ΔBMI and blood tissue

| **Ethnicity** | **Ensembl_Gene_ID** | **Chr** | **Gene** | **Top_SNP** | **p_SMR** | **p_HEIDI** | **nSNP_HEIDI** |
| --- | --- | --- | --- | --- | --- | --- | --- |
| EUR | ENSG00000101558 | 18 | *VAPA* | rs624898 | 6.60E-04 | 6.76E-01 | 7 |
|  | ENSG00000179979 | 4 | *CRIPAK* | rs1250121 | 1.26E-03 | 1.23E-01 | 13 |
|  | ENSG00000204713 | 6 | *TRIM27* | rs4713186 | 1.27E-03 | 4.06E-01 | 20 |
|  | ENSG00000184361 | 17 | *SPATA32* | rs4986168 | 1.96E-03 | 1.25E-01 | 8 |
|  | ENSG00000182240 | 21 | *BACE2* | rs914178 | 2.19E-03 | *NA* | *NA* |
|  | ENSG00000067836 | 16 | *ROGDI* | rs2063101 | 2.32E-03 | 5.04E-01 | 8 |
|  | ENSG00000112531 | 6 | *QKI* | rs2759386 | 2.97E-03 | *NA* | *NA* |
|  | ENSG00000118514 | 6 | *ALDH8A1* | rs1014021 | 3.99E-03 | 9.02E-01 | 7 |
|  | ENSG00000110025 | 11 | *SNX15* | rs502333 | 4.08E-03 | 4.84E-02 | 3 |
|  | ENSG00000267077 | 16 | *RP11-127I20.5* | rs8064024 | 4.54E-03 | 8.93E-01 | 11 |
|  | ENSG00000235169 | 1 | *SMIM1* | rs10797348 | 4.98E-03 | 3.88E-01 | 8 |
| AFR | ENSG00000138030 | 2 | *KHK* | rs3769139 | 9.83E-04 | 8.97E-01 | 20 |
|  | ENSG00000173578 | 3 | *XCR1* | rs9311384 | 1.04E-03 | 3.96E-01 | 6 |
|  | ENSG00000260135 | 16 | *RP11-212I21.2* | rs1345319 | 1.77E-03 | 1.46E-01 | 3 |
|  | ENSG00000254614 | 11 | *AP003068.23* | rs1195968 | 2.14E-03 | 2.35E-01 | 13 |
|  | ENSG00000165030 | 9 | *NFIL3* | rs1751796 | 2.49E-03 | 6.16E-02 | 7 |
|  | ENSG00000229515 | 3 | *FLT1P1* | rs4490403 | 2.66E-03 | 4.78E-01 | 4 |
| HIS | ENSG00000183323 | 5 | *CCDC125* | rs12513817 | 1.03E-03 | 4.77E-01 | 8 |
|  | ENSG00000168060 | 11 | *NAALADL1* | rs3896078 | 1.85E-03 | 6.02E-01 | 5 |
|  | ENSG00000167491 | 19 | *GATAD2A* | rs2965180 | 1.94E-03 | 7.43E-01 | 12 |
|  | ENSG00000135951 | 2 | *TSGA10* | rs6727749 | 1.98E-03 | 5.42E-01 | 15 |
|  | ENSG00000144182 | 2 | *LIPT1* | rs4851188 | 2.10E-03 | 5.69E-01 | 10 |
|  | ENSG00000127948 | 7 | *POR* | rs10954732 | 2.68E-03 | 8.66E-01 | 6 |
|  | ENSG00000141219 | 17 | *C17orf80* | rs3751925 | 3.35E-03 | 7.59E-01 | 7 |
|  | ENSG00000160113 | 19 | *NR2F6* | rs8108035 | 3.38E-03 | *NA* | *NA* |
|  | ENSG00000159388 | 1 | *BTG2* | rs4971226 | 3.64E-03 | 8.51E-01 | 3 |
|  | ENSG00000105793 | 7 | *GTPBP10* | rs177665 | 3.91E-03 | 1.66E-01 | 10 |
|  | ENSG00000198885 | 2 | *ITPRIPL1* | rs772174 | 4.26E-03 | 6.28E-01 | 13 |
|  | ENSG00000273045 | 2 | *C2ORF15* | rs12613624 | 4.43E-03 | 6.76E-01 | 8 |
|  | ENSG00000120675 | 13 | *DNAJC15* | rs4942185 | 4.44E-03 | 2.30E-01 | 12 |
|  | ENSG00000182919 | 11 | *C11orf54* | rs1284108 | 4.66E-03 | 8.70E-01 | 8 |
|  | ENSG00000213830 | 5 | *CFL1P5* | rs350104 | 4.71E-03 | 4.10E-01 | 7 |
|  | ENSG00000182534 | 17 | *MXRA7* | rs9915664 | 4.72E-03 | 1.60E-01 | 8 |
|  | ENSG00000239672 | 17 | *NME1* | rs3760469 | 4.98E-03 | 9.01E-01 | 8 |

Table S12. SMR results for ΔBMI and liver tissue

| **Ethnicity** | **Ensembl_Gene_ID** | **Chr** | **Gene** | **Top_SNP** | **p_SMR** | **p_HEIDI** | **nSNP_HEIDI** |
| --- | --- | --- | --- | --- | --- | --- | --- |
| EUR | ENSG00000140623 | 16 | *SEPT12* | rs9925944 | 1.87E-03 | 6.40E-01 | 4 |
|  | ENSG00000227258 | 13 | *SMIM2-AS1* | rs9525917 | 4.92E-03 | 5.58E-01 | 5 |
| AFR | ENSG00000140367 | 15 | *UBE2Q2* | rs4886753 | 1.92E-03 | *NA* | *NA* |
| HIS | ENSG00000271553 | 7 | *RP11-274B21.10* | rs339072 | 1.55E-03 | 2.89E-01 | 3 |
|  | ENSG00000135951 | 2 | *TSGA10* | rs6727749 | 2.87E-03 | 6.12E-01 | 8 |
|  | ENSG00000267594 | 19 | *CYP4F24P* | rs10419282 | 4.34E-03 | 3.23E-01 | 3 |
|  | ENSG00000144182 | 2 | *LIPT1* | rs6727749 | 4.40E-03 | 7.24E-01 | 7 |

Table S13. SMR results for ΔBMI > 1.5 kg/m² and blood tissue

| **Ethnicity** | **Ensembl_Gene_ID** | **Chr** | **Gene** | **Top_SNP** | **p_SMR** | **p_HEIDI** | **nSNP_HEIDI** |
| --- | --- | --- | --- | --- | --- | --- | --- |
| EUR | ENSG00000122873 | 10 | *CISD1* | rs1698469 | 2.91E-04 | 7.40E-02 | 4 |
|  | ENSG00000133466 | 22 | *C1QTNF6* | rs4821603 | 9.62E-04 | *NA* | *NA* |
|  | ENSG00000257497 | 12 | *RP11-585P4.5* | rs12830818 | 1.45E-03 | 6.36E-01 | 11 |
|  | ENSG00000101558 | 18 | *VAPA* | rs624898 | 1.85E-03 | 4.30E-01 | 7 |
|  | ENSG00000055950 | 10 | *MRPL43* | rs11595160 | 2.25E-03 | 2.87E-01 | 3 |
|  | ENSG00000140575 | 15 | *IQGAP1* | rs2074585 | 2.31E-03 | 9.40E-01 | 7 |
|  | ENSG00000189060 | 22 | *H1F0* | rs5756825 | 2.74E-03 | 3.55E-01 | 19 |
|  | ENSG00000197147 | 1 | *LRRC8B* | rs6699344 | 2.85E-03 | 4.89E-01 | 14 |
|  | ENSG00000134248 | 1 | *LAMTOR5* | rs6671673 | 2.91E-03 | 8.51E-01 | 8 |
|  | ENSG00000214309 | 7 | *MBLAC1* | rs4729565 | 2.97E-03 | 5.37E-01 | 3 |
|  | ENSG00000246228 | 8 | *CASC8* | rs7012462 | 3.03E-03 | 8.79E-02 | 5 |
|  | ENSG00000235169 | 1 | *SMIM1* | rs10797348 | 3.40E-03 | 4.57E-01 | 8 |
|  | ENSG00000168090 | 7 | *COPS6* | rs4729565 | 3.77E-03 | 3.89E-01 | 3 |
|  | ENSG00000154124 | 5 | *FAM105B* | rs25986 | 4.11E-03 | 5.29E-01 | 8 |
|  | ENSG00000215267 | 10 | *AKR1C7P* | rs2151896 | 4.51E-03 | *NA* | *NA* |
| AFR | ENSG00000132436 | 7 | *FIGNL1* | rs11768267 | 6.71E-04 | 1.97E-02 | 20 |
|  | ENSG00000089902 | 14 | *RCOR1* | rs4906239 | 3.41E-03 | 8.24E-01 | 3 |
|  | ENSG00000167207 | 16 | *NOD2* | rs1981760 | 3.50E-03 | 4.37E-01 | 8 |
|  | ENSG00000171135 | 3 | *JAGN1* | rs279542 | 3.67E-03 | 9.36E-02 | 16 |
|  | ENSG00000095321 | 9 | *CRAT* | rs7855278 | 3.83E-03 | 4.23E-01 | 12 |
|  | ENSG00000270120 | 16 | *RP11-327F22.6* | rs1981760 | 3.88E-03 | 4.61E-01 | 8 |
|  | ENSG00000119383 | 9 | *PPP2R4* | rs7855278 | 3.94E-03 | 4.68E-01 | 12 |
|  | ENSG00000267769 | 19 | *CTB-50L17.9* | rs243372 | 4.30E-03 | 9.19E-01 | 10 |
|  | ENSG00000129295 | 8 | *LRRC6* | rs1895807 | 4.36E-03 | 2.83E-01 | 11 |
|  | ENSG00000230732 | 3 | *AC127904.2* | rs9784336 | 4.49E-03 | 3.47E-02 | 9 |
|  | ENSG00000112110 | 6 | *MRPL18* | rs2273827 | 4.52E-03 | 7.23E-01 | 12 |
|  | ENSG00000163702 | 3 | *IL17RC* | rs708567 | 4.64E-03 | 1.17E-01 | 16 |
|  | ENSG00000211456 | 3 | *SACM1L* | rs1962800 | 4.68E-03 | 2.68E-02 | 4 |
|  | ENSG00000224152 | 2 | *AC009506.1* | rs6743189 | 4.75E-03 | 3.44E-01 | 5 |
| HIS | ENSG00000145220 | 4 | *LYAR* | rs2920174 | 4.90E-04 | 6.37E-01 | 4 |
|  | ENSG00000134287 | 12 | *ARF3* | rs833830 | 1.58E-03 | *NA* | *NA* |
|  | ENSG00000148110 | 9 | *HIATL1* | rs2769816 | 3.00E-03 | 9.98E-01 | 5 |
|  | ENSG00000025156 | 6 | *HSF2* | rs4945703 | 3.17E-03 | 4.28E-01 | 20 |
|  | ENSG00000239665 | 10 | *RP11-295P9.3* | rs4500387 | 3.28E-03 | 8.25E-01 | 11 |
|  | ENSG00000242616 | 9 | *GNG10* | rs7045665 | 3.50E-03 | 5.70E-02 | 14 |
|  | ENSG00000125810 | 20 | *CD93* | rs844887 | 3.59E-03 | 7.06E-01 | 20 |
|  | ENSG00000107331 | 9 | *ABCA2* | rs4880192 | 4.53E-03 | 6.33E-01 | 8 |

Table S14. SMR results for ΔBMI > 1.5 kg/m² and liver tissue

| **Ethnicity** | **Ensembl_Gene_ID** | **Chr** | **Gene** | **Top_SNP** | **p_SMR** | **p_HEIDI** | **nSNP_HEIDI** |
| --- | --- | --- | --- | --- | --- | --- | --- |
| EUR | ENSG00000257497 | 12 | *RP11-585P4.5* | rs7300308 | 1.24E-03 | 5.94E-01 | 3 |
|  | ENSG00000180481 | 12 | *GLIPR1L2* | rs12830818 | 1.73E-03 | 9.05E-01 | 3 |
|  | ENSG00000133466 | 22 | *C1QTNF6* | rs4821603 | 2.40E-03 | *NA* | *NA* |
| AFR | ENSG00000243317 | 7 | *C7orf73* | rs6467595 | 9.70E-04 | 1.22E-03 | 3 |
|  | ENSG00000250786 | 5 | *SNHG18* | rs218057 | 1.62E-03 | *NA* | *NA* |
|  | ENSG00000150967 | 12 | *ABCB9* | rs1727307 | 2.30E-03 | 3.51E-01 | 12 |
|  | ENSG00000105755 | 19 | *ETHE1* | rs2854506 | 3.16E-03 | 6.35E-01 | 9 |
|  | ENSG00000198829 | 3 | *SUCNR1* | rs6440788 | 3.39E-03 | *NA* | *NA* |
| HIS | ENSG00000242616 | 9 | *GNG10* | rs7045665 | 4.54E-03 | 1.48E-01 | 13 |
|  | ENSG00000271553 | 7 | *RP11-274B21.10* | rs339072 | 4.73E-03 | 6.49E-01 | 3 |
