## Supplemental File 2 for "Genomic and transcriptomic insights into antipsychotic-induced changes in total cholesterol and body mass index in a multi-ancestry cohort of the US veterans"

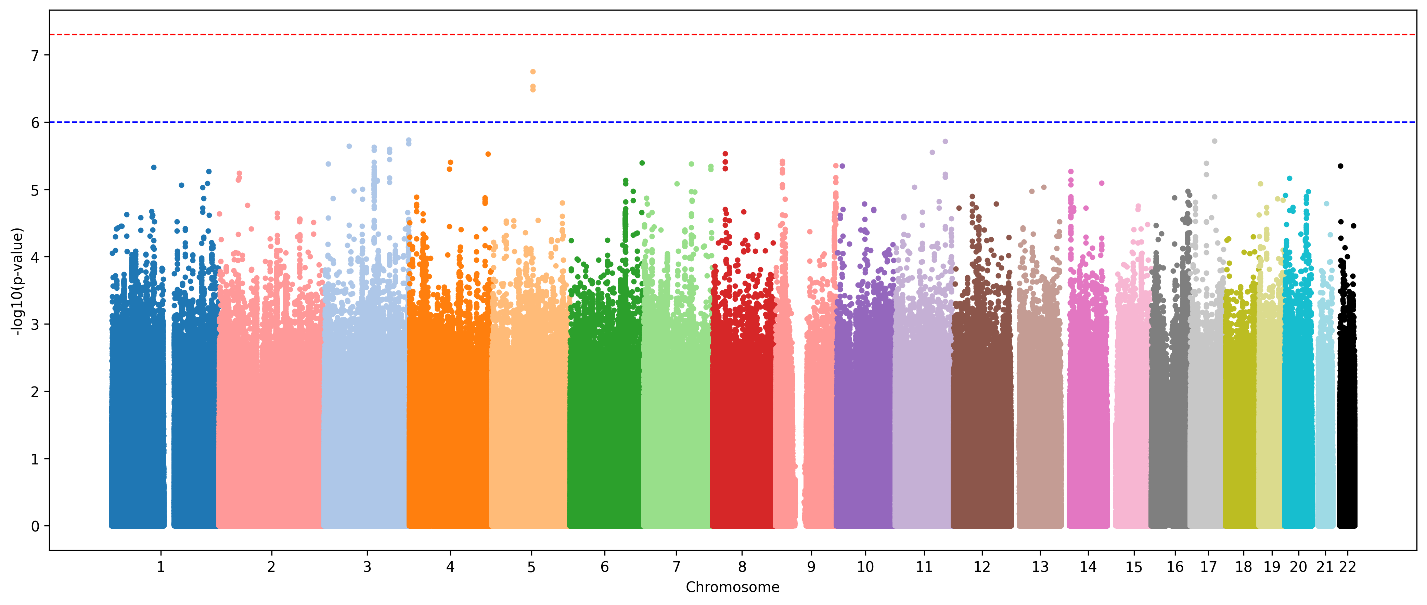
Figure S5. Manhattan plot of GWAS results for ΔTC in HIS cohort

**----** Genome-wide line

(p = 5×10^-8^)

**----** Suggestive line

(p = 1×10^-6^)

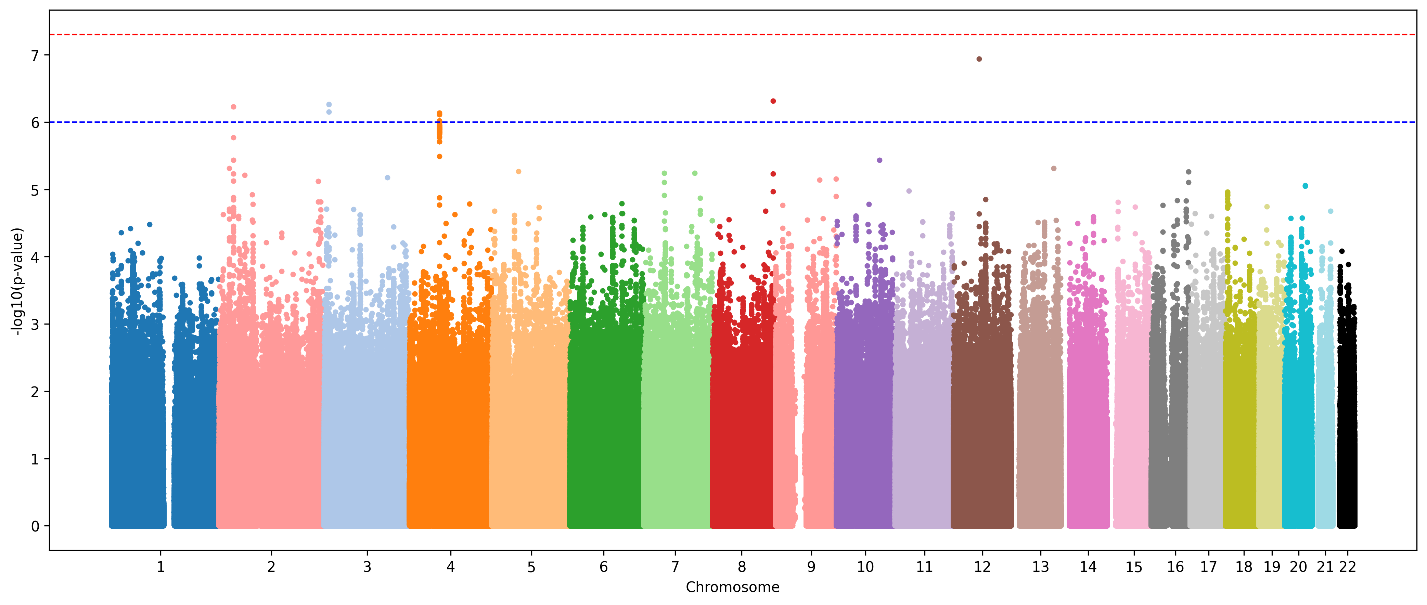
Figure S6. Manhattan plot of GWAS results for ΔBMI in EUR cohort

**----** Genome-wide line

(p = 5×10^-8^)

**----** Suggestive line

(p = 1×10^-6^)

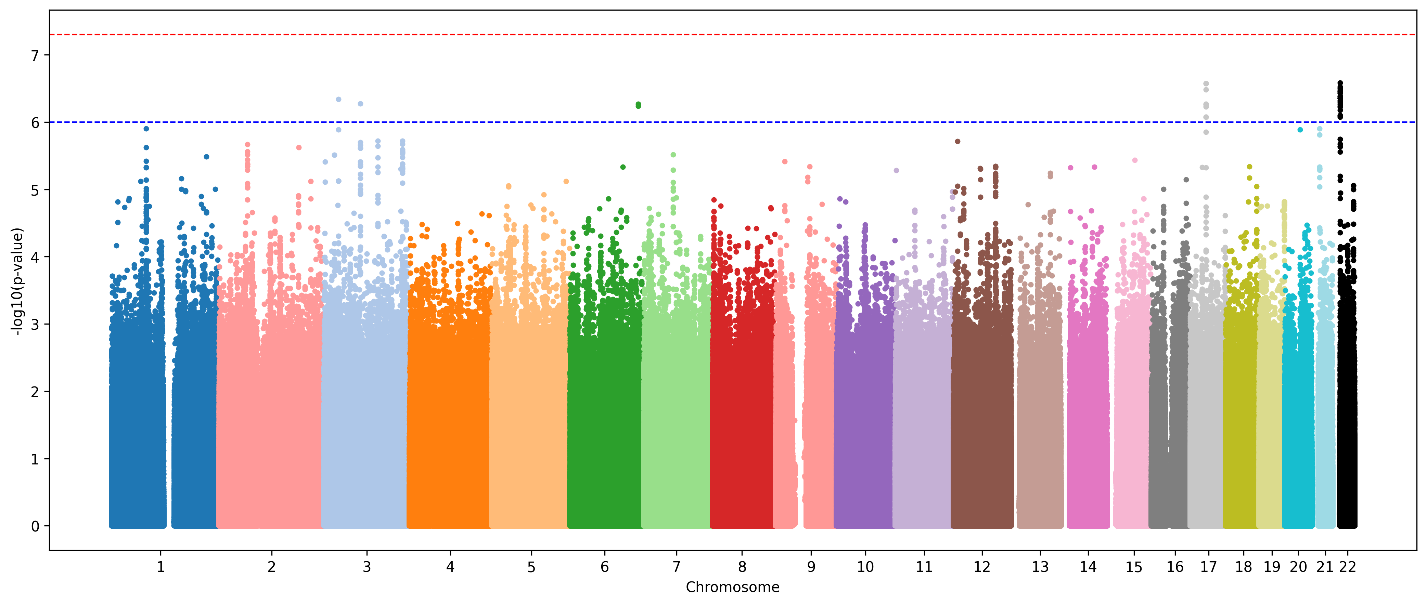
Figure S7. Manhattan plot of GWAS results for ΔBMI in AFR cohort

**----** Genome-wide line

(p = 5×10^-8^)

**----** Suggestive line

(p = 1×10^-6^)

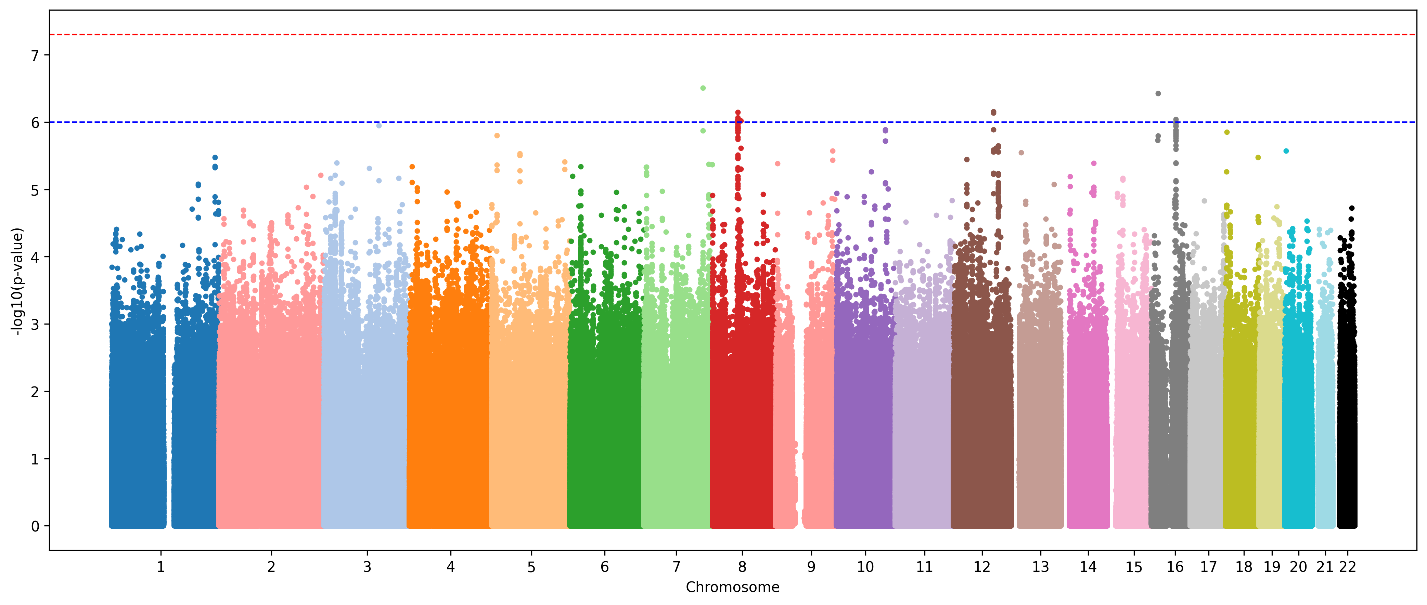
Figure S8. Manhattan plot of GWAS results for ΔBMI in HIS cohort

**----** Genome-wide line

(p = 5×10^-8^)

**----** Suggestive line

(p = 1×10^-6^)

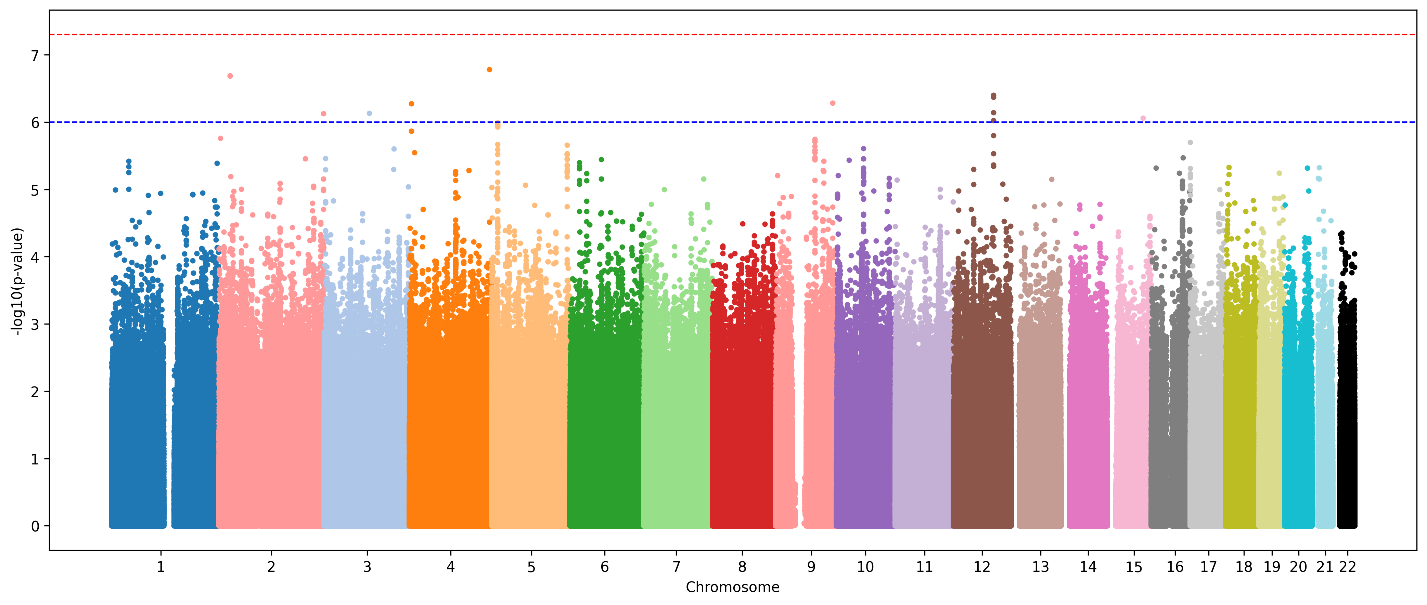
Figure S9. Manhattan plot of GWAS results for ΔBMI > 1.5 kg/m² in HIS cohort

**----** Genome-wide line

(p = 5×10^-8^)

**----** Suggestive line

(p = 1×10^-6^)

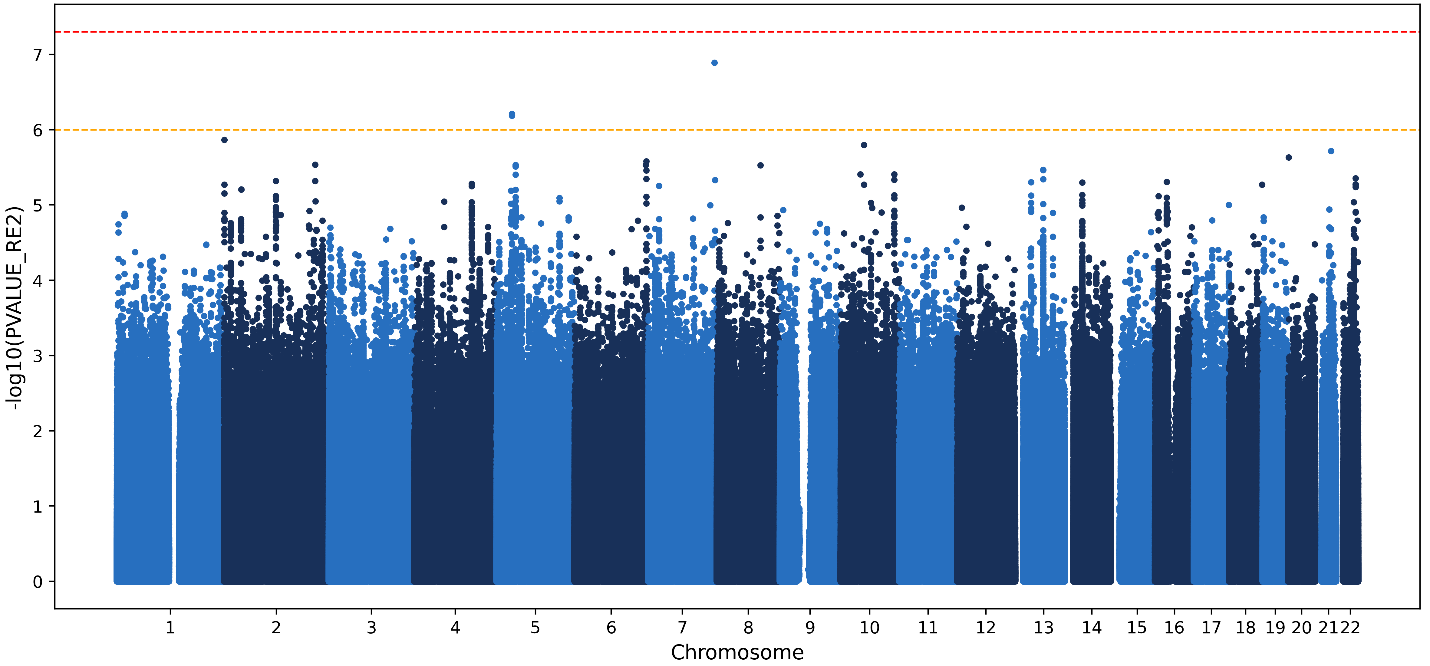
Figure S10. Manhattan plot of meta-analysis of GWAS results for ΔTC

*DPP6*

**----** Genome-wide line

(p = 5×10^-8^)

**----** Suggestive line

(p = 1×10^-6^)

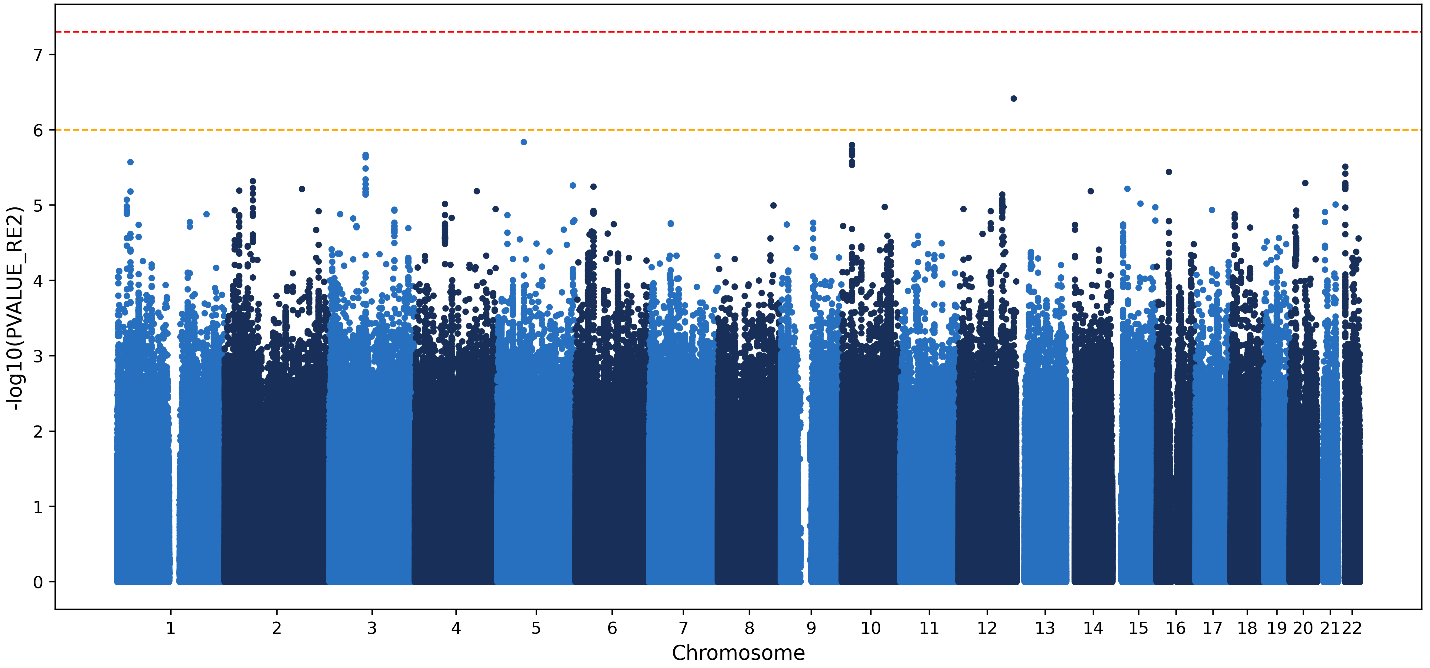
Figure S11. Manhattan plot of meta-analysis of GWAS results for ΔBMI

**----** Genome-wide line

(p = 5×10^-8^)

**----** Suggestive line

(p = 1×10^-6^)

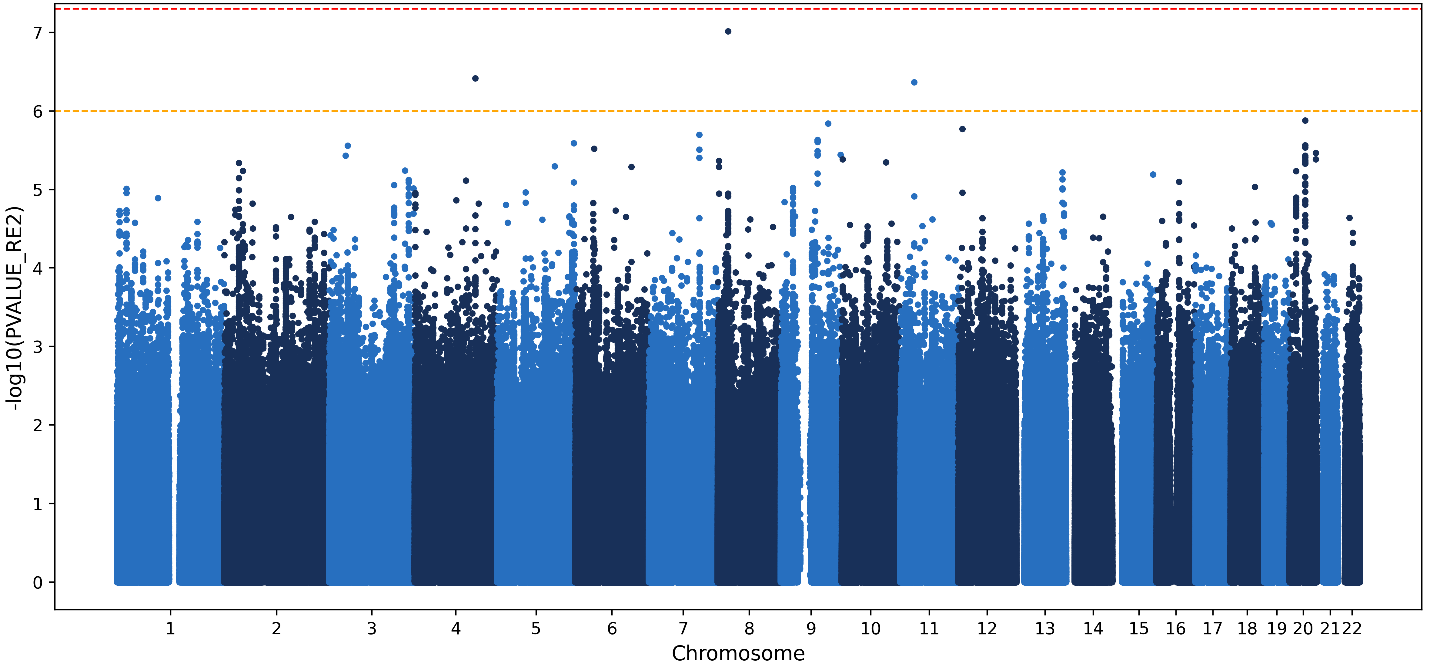
Figure S12. Manhattan plot of meta-analysis of GWAS results for ΔBMI > 1.5 kg/m²

**----** Genome-wide line

(p = 5×10^-8^)

**----** Suggestive line

(p = 1×10^-6^)

*ADAM28, ADAMDEC1*
